# Optimizing Gene Selection and Network-Level Insights in Hypertrophic Cardiomyopathy: A Novel Genetic Algorithm Combined with WGCNA and Statistical Filtering

**DOI:** 10.1101/2025.07.01.25330641

**Authors:** Supriya Mandal, Apurvanand Sahaya, Amrita Thakur, Surama Biswas

## Abstract

A cardiac condition known as hypertrophic cardiomyopathy (HCM) is characterized by an irregular thickening of the heart muscle. There is still much to learn about its molecular mechanics. In order to pinpoint important genes and regulatory abnormalities in HCM, this work offers a thorough computational analysis of gene expression data. Two strategies are employed here. Initially, hub genes were identified, co-expression networks were constructed, gene modules were detected, and they were linked to clinical characteristics using Weighted Gene Co-expression Network Analysis (WGCNA). Second, the same dataset was subjected to three different gene selection techniques: variance-based filtering, volcano plot analysis, and a Genetic Algorithm for Novel Gene Acquisition (GANGA). For the first time, GANGA successfully incorporates a previously defined objective function from simulated annealing into a genetic algorithm. Additionally, it uses two-point crossover, meticulous parameter optimization, and customizable elitism. Three genes were shown to be shared by all approaches, including WGCNA: RASID1, CEBPD, and S100A9. Through enrichment analysis, these were confirmed to be implicated in pathways linked to inflammation. Their incorporation into cytokine-driven networks was validated by investigation of protein-protein interactions. S100A9 emerged as a crucial regulator that activates RASD1 in illness, according to co-expression networks constructed for normal and HCM samples, which showed changed regulatory patterns. The methodological development of modifying and optimizing a simulated annealing-based objective function within a GA framework in GANGA for efficient gene selection, as well as the comprehensive multi-method pipeline for HCM analysis, are what make this study distinctive.

## 1. INTRODUCTION

With significant mortality and morbidity, cardiomyopathy—which is associated with cardiac muscle or electrical abnormalities of the heart—often results in progressive heart failure. The three main variants are hypertrophic cardiomyopathy (HCM), dilated cardiomyopathy (DCM), and restricted cardiomyopathy (RCM). It is divided into primary (inherited and/or acquired) and secondary (inflammatory and/or toxic) types (Wexler et. al., 2009). About 1 in 500 people worldwide suffer with HCM, the most prevalent genetic cardiomyopathy, which is a major cause of sudden cardiac mortality, particularly in sports and young people. Because of its serious implications, HCM is frequently underdiagnosed, which can raise mortality. Although its penetrance and expression might vary, HCM is primarily a single-gene illness with an autosomal dominant inheritance pattern, meaning that a single mutation can produce the disease. Rare autosomal recessive and X-linked inheritance patterns are also present, however familial inheritance accounts for about 60% of HCM cases. Research has shown that more than 1500 mutations in the genes encoding cardiac sarcomere proteins cause the disorder (Varma and Neema, 2014; Varma et. al., 2015; Chen et al., 2024).

Myocyte hypertrophy, interstitial fibrosis, and disarray are among the histological markers of the disease, which can lead to left ventricular diastolic failure. The disease also has a complex pathophysiology and a diverse clinical presentation. Left ventricular hypertrophy, systolic anterior motion of the mitral valve, blockage of the left ventricular outflow tract (LVOT), mitral regurgitation, and myocardial ischemia are pathological features of HCM. Heart failure and strokes brought on by atrial fibrillation are common complications. Serum chemical testing, echocardiography, and electrocardiography are commonly used to diagnose the condition. Through heart transplantation, cardiac resynchronization therapy, and medication, as well as lifestyle changes like cutting back on alcohol and sodium, losing weight, exercising, and giving up smoking, treatment strategies seek to lower hospitalization and mortality rates (Tuohy et al., 2020). Current treatments include medications, myectomy, alcohol septal ablation, implanted cardioverter defibrillators (ICDs), and other procedures that focus on managing symptoms and delaying the condition’s progression in order to prevent sudden cardiac death. For those with severe symptoms, invasive therapies may be required, even though medication is the basis of treatment. Despite major improvements in care, HCM remains a serious burden, even with the opportunity for new treatments offered by innovative pharmacotherapies, minimally invasive procedures, and gene-directed approaches. Patients with obstructive HCM may have options thanks to novel surgical approaches that could eventually address the genetic abnormalities causing the condition. Though they have mostly failed, recent pharmacological studies have helped us better understand therapeutic targets (Maron et al., 2022).

The knowledge of molecular components has advanced relatively slowly, despite significant improvements in clinical practice. In the past few decades, molecular genetic insights into primary cardiomyopathies, including familial hypertrophic and X-linked dilated cardiomyopathy, have surfaced alongside ongoing research on other heart disorders TOwBIN (1993). Pathogenic mutations in genes that cause inherited cardiac arrhythmias have been connected to some cases of hereditary cardiomyopathy, such as HCM, suggesting a shared genetic origin. However, the precise etiology of these genes remains unknown. In hereditary cardiomyopathies, arrhythmias are caused by structural changes such as hypertrophy and fibrosis as well as changes in secondary ion currents. Thanks to developments in molecular genetics, more than a dozen causal genes have been identified; mutations in MYH7 and MYBPC3 are responsible for 50% of cases. Genetic screening is now possible, however it is still challenging to distinguish between causative and non-causal variants. Effective treatment requires an understanding of the various processes behind HCM. (Marian, 2010; Masarone et al., 2018; Bayes-Genis et al., 2020; Crotti et al., 2023; Asatryan et al. 2024). According to Biswas et al., 2022, a deeper understanding of the metabolism pathways is necessary in order to discover the genes implicated in the processes that result in drug resistance.

One popular technique for examining gene correlation patterns in systems biology is WGCNA, introduced by Langfelder and Horvath, (2008). Modules are collections of highly co-expressed genes that are identified using WGCNA. Hub genes or module eigengenes can be used to summarize these. After the modules are connected to external characteristics, WGCNA can be used to identify treatment targets and find biomarkers. DiLeo et al. (2011) used WGCNA in metabolomics to investigate tomato fruit development. Modules of co-expressed metabolites linked to genetic background and ripening were discovered. Zhu et al. (2018) discovered modules linked to cell cycle control and oxidative metabolism in hepatocellular carcinoma (HCC). Clinical stages of HCC were associated with these modules (Yin et al., 2018). Four hub genes—FAM171A1, NDFIP1, SKP1, and REEP5— were identified by Tian et al. (2020) as possible biomarkers for individualized treatment in breast cancer (BRCA).

WGCNA and machine learning (ML) together improve the identification of genes linked to disease and intricate biological processes. Gene modules linked to clinical characteristics are identified by WGCNA (Chen et al., 2022). Within these modules, gene selection is refined by machine learning techniques such as random forests, SVM-RFE, and LASSO regression. When combined, WGCNA and machine learning (ML) enhance the detection of genes associated with complex biological processes and disease. WGCNA identifies gene modules associated with clinical features. Machine learning methods including random forests, SVM-RFE, and LASSO regression are used in these modules to improve gene selection. Zhao et al. (2022) developed a predictive model that forecasted immunological function and patient outcomes in ovarian cancer using glycosylation-related mRNAs. Xu et al. (2023) discovered hub genes (CD36, ITGB2, SLC1A3) connected to immunity and oxidative stress in diabetic nephropathy. This method was used more recently in ischemic cardiomyopathy and IgA nephropathy by Kong et al. (2023) and Ren et al. (2024), who discovered novel biomarkers and patterns of immune cell infiltration. These results validate the effectiveness of integrating machine learning and WGCNA in the identification of disease genes.

## 2. RELATED WORKS

Finding biomarkers for different cardiomyopathies, such as hypertrophic (HCM), dilated (DCM), and ischemic (ICM) cardiomyopathies, has been the focus of recent research. For example, it was discovered that DCM patients had elevated levels of NPPA, OMD, and PRELP, which were associated with the TGF-β signaling pathway (Cao et al., 2021). Immune infiltration and mitochondrial damage were linked to genes such as CASP3 and CHCHD4 in ICM-HF (Zhu et al., 2022). Genes such as COL3A1 and LUM in ICM have also been validated by research (Yuan et al., 2024a). These biomarkers improve our knowledge of the development of cardiomyopathy, particularly as it relates to extracellular matrix remodeling and immune response. Strong gene-level indicators for HCM that have been confirmed by both co-expression and enrichment analysis have, however, not been thoroughly investigated (Wang et al., 2022; Yuan et al., 2024b).

WGCNA has been used extensively to find hub genes and gene modules linked to cardiovascular disorders. Candidate genes such as AQP3 and CYP2J2 were chosen from modules associated with extracellular matrix function that were identified by WGCNA in IDCM-HF (Li et al., 2024). Similar methods were used in DCM and ICM to identify functional modules and hub genes related to fibrosis and immunological responses. PPI networks and enrichment analysis were used in these investigations to confirm biological significance. WGCNA’s use in HCM is still limited, despite its critical role in finding trait-associated modules. This is particularly true when comparing healthy and sick circumstances using integrated network and pathway-level insights (Wang et al., 2019).

WGCNA and AIML models work together to increase biomarker detection accuracy. Gene sets chosen from WGCNA modules have been successfully refined in studies employing LASSO, SVM-RFE, and random forests (Yu et al., 2016). For instance, using integrated analysis, four genes—IL1B, TIMP2, IFIT3, and P2RY2—were shown to be involved in IHF and confirmed to have an AUC > 0.9 (Dang et al., 2020). Even though these integrated approaches show promise, they are rarely investigated in the context of HCM, particularly when compared to co-expression changes and pathway-level dynamics (Yu et al., 2023). According to Li et al. (2024), ML assisted in the selection of NPPA and CYP2J2 with high diagnostic potential in DCM. Additionally, these hybrid techniques have revealed patterns of immune cell infiltration.

Finding gene subsets that best distinguish between healthy and diseased samples can be done by modeling gene selection as an optimization problem. In this field, meta-heuristic techniques such as Genetic Algorithms (GA) and Simulated Annealing (SA) have demonstrated potential. Differential Gene Expression Based Simulated Annealing (DGESA), as proposed by Sinha et al. (2024), ranked gene sets according to their combined capacity to differentiate expression profiles. DGESA found a number of verified genes, such as SPRR2B and STAT6, that are supported by literature and enrichment analysis when applied to Eosinophilic Esophagitis (EoE) and other gastrointestinal disorders. These biologically based optimization techniques supplement conventional statistical methods in gene expression research and improve the identification of disease genes.

HCM-specific research is still underrepresented, despite the fact that biomarker discovery for cardiomyopathies utilizing WGCNA and ML is well-documented. There aren’t many integrated approaches in the literature that include co-expression modeling, meta-heuristic optimization, robust statistical filters, and pathway-level validation in a single pipeline. There aren’t many ways to compare the effectiveness of different gene selection techniques or confirm the findings using network dynamics and enriched biological functions. Our study fills this gap by integrating WGCNA, variance and Volcano plot filtering, and Genetic Algorithm for Novel Gene Acquisition (GANGA), a novel GA-based gene selector, and to create a coherent analysis of HCM gene regulation. The goal of this integrated strategy is to reconcile biological interpretability with computational rigor.

## 3. CONTRIBUTIONS

### 3.1. Context and Objective

Abnormal thickening of the heart muscle is a hallmark of hypertrophic cardiomyopathy (HCM), a severe cardiac disorder that frequently results in major problems and increased health risks. Identifying diagnostic and treatment targets requires an understanding of the molecular basis of HCM. Finding important regulatory genes and clarifying the related biological mechanisms are the main goals of this study’s thorough gene expression investigation of HCM. Our approach combines targeted gene selection methods with co-expression network modeling to do this, concentrating on both the statistical differences in gene expression between healthy and ill individuals as well as the structural patterns within gene networks.

### 3.2. Methodological innovation

We build scale-free gene networks, identify hub genes of interest, detect co-expressed modules, and link them with external attributes using WGCNA. We simultaneously use the same dataset to test three machine learning and statistical methods: a novel genetic algorithm, variance-based selection, and volcano plot analysis. GANGA, our suggested genetic algorithm, incorporates biologically significant limitations like elitism, non-redundant chromosomes, and a unique two-point crossover. The objective function was first presented in our earlier work, but this is the first time it has been included into a new GA-based architecture.

### 3.3. Results and biological insight

We discover important gene subsets with overlapping members by using statistical techniques supported by both WGCNA and GANGA to analyze the HCM dataset. Ten common genes, such as SERPINE1, S100A9, and CORIN, are consistently picked using variance, volcano, and GA techniques without taking WGCNA into account. Three overlapping genes—RASD1, CEBPD, and S100A9—emerge when WGCNA is incorporated into the study, underscoring the extra knowledge obtained from network-based gene prioritization. Through the use of Metascape’s enrichment and protein interaction analyses, these genes were confirmed to have close ties to cytokine-regulated and inflammatory pathways. Co-expression networks for healthy and sick states also showed changes in regulation, especially the part S100A9 plays in RASD1 activation in the illness.

### 3.4. Novelty and claim

This research contributes in two ways. It starts by presenting GANGA, a specially designed genetic method for transcriptomic study gene selection. GANGA reuses a previously established goal function while incorporating new evolutionary mechanisms, such as physiologically aware two-point crossover, elitism, and gene-level uniqueness enforcement that are optimized for high-dimensional omics data. Second, by combining GANGA with WGCNA and additional selection techniques, the study finds genetic markers for Hypertrophic Cardiomyopathy (HCM) that are relevant to the condition. Notably, major overlapping genes across techniques and network studies were found to include S100A9, RASD1, and CEBPD. These genes showed different regulatory functions between healthy and sick co-expression networks and were enriched in pathways associated to inflammation and cytokines, providing new information about the molecular basis of HCM.

## 4. METHODOLOGIES

The analytical methodology used to pinpoint important genes and molecular pathways linked to hypertrophic cardiomyopathy (HCM) is described in this section. Variance filtering, volcano plot analysis, and a customized Genetic Algorithm, GANGA are used for gene selection after the preprocessing of gene expression data. Hub genes are found and gene modules are constructed using WGCNA. In order to identify consistently relevant biomarkers, gene overlaps across all selection approaches are subsequently analyzed. To find structural differences, co-expression networks are created and analyzed for samples that are healthy and samples that are ill. Lastly, protein–protein interaction (PPI) network creation and pathway enrichment analysis are used to confirm the biological importance of the chosen genes (see Figure 1).

**Figure 1.**
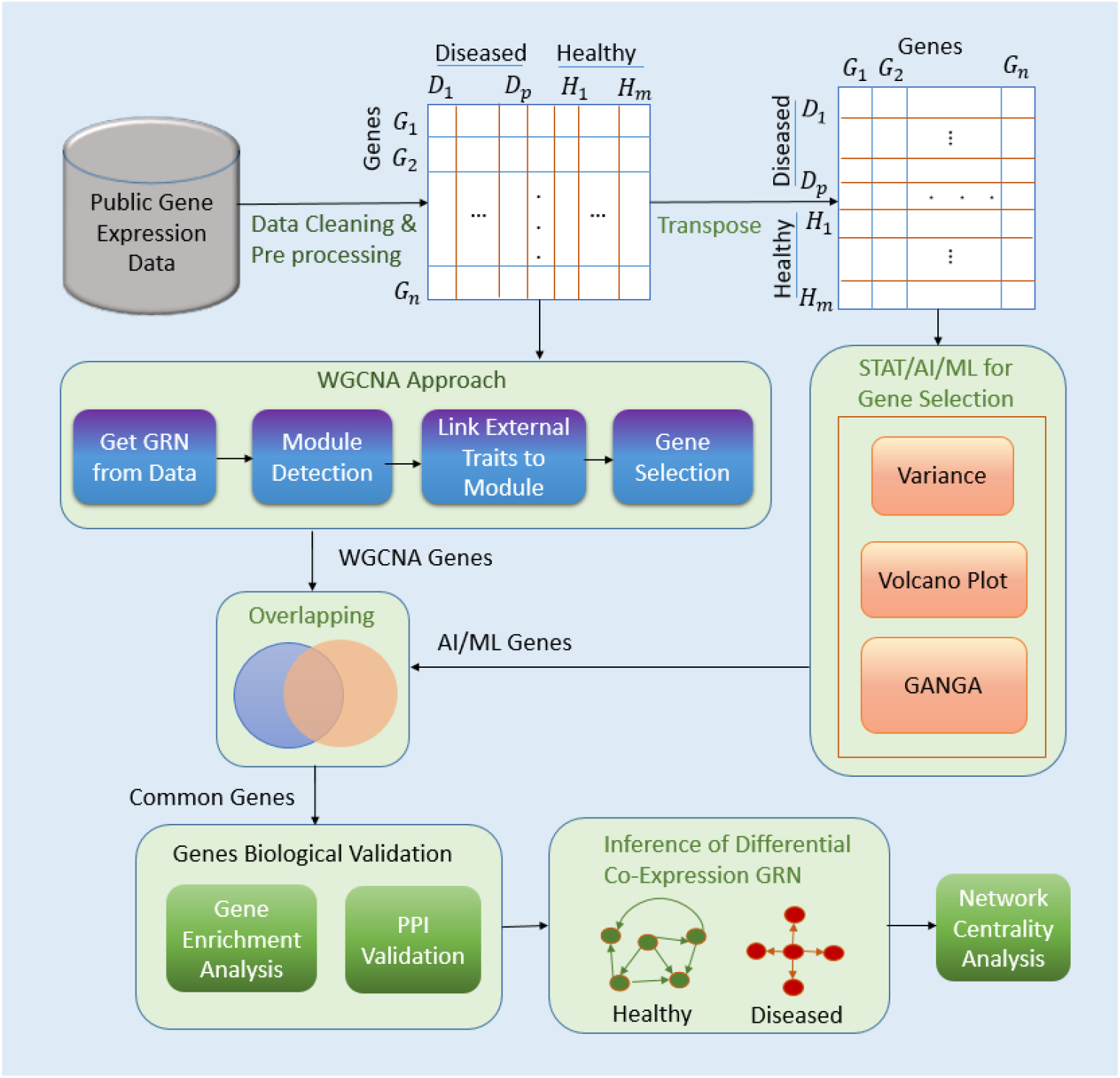
An outline of the suggested analytical framework for the analysis of HCM genes. Preprocessing gene expression data, gene selection using WGCNA, variance, volcano plot, and GANGA techniques, finding overlapping genes, creating co-expression networks for both healthy and diseased states, and downstream validation using enrichment and PPI network analyses are all steps in the pipeline.

### 4.1. Data Acquisition and Preprocessing Strategy

The study begins with the collection of a gene expression matrix, where the rows represent individual genes and the columns correspond to different samples. Before analysis, thorough quality control is performed to ensure the integrity of the data. This involves the elimination of genes or samples that exhibit poor quality, have excessive missing data, or show insufficient variability. Such preprocessing steps are critical to maintaining the reliability of subsequent analyses. Following quality control, the data undergo normalization to transform the gene expression levels, enabling comparability across samples. Normalization is essential for adjusting systematic biases and ensuring that observed differences in expression levels accurately reflect biological variations rather than technical artifacts. These preprocessing steps lay the foundation for robust data analysis, enhancing the accuracy of the insights gained from the subsequent gene expression analysis.

### 4.2. Analysis of Gene Expression Patterns in HCM through WGCNA

WGCNA refers to the system of biological methods applied for describing the correlation patterns among genes across many samples. It is useful in identifying clusters, or modules, of highly correlated genes, and summarizing these modules with module eigengenes, and relating them to external traits or conditions (Langfelder and Horvath, 2008). WGCNA also provides tools suitable for network-based gene screening methods that identify candidate biomarkers or therapeutic targets. The detailed steps of WGCNA are explained below.

#### 4.2.1.Construction of a Gene Co-expression Network

The main goal of this part of the work is to convert the gene expression matrix into a weighted co-expression network, in which each node stands for a gene and the edges between nodes indicate how strongly those nodes co-express. This is accomplished by comparing the expression profiles of genes across samples to determine how similar they are. Because it helps identify linear correlations between gene expression patterns, Pearson correlation (Eq. 1) is the most widely used similarity metric.

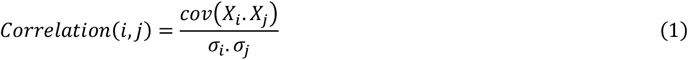

Here, *X*_*i*_ and *X*_*j*_ Are the vectors of the expression for the genes *i* and *j*, with standard deviations σ_*i*_ and. σ_*j*_ respectively. To achieve a scale-free topology (in the form of an adjacency matrix, *A*(*i, j*)) typical of biological networks, the absolute correlation is raised to a power β after computing similarity (see Eq. 2).

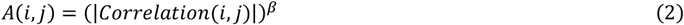

Using a power transformation in WGCNA increases stronger correlations and decreases weaker ones. A soft-thresholding power β is used for this. The best fit for a scale-free topology is determined by visually examining plots to determine the value of β. Few genes have numerous links in these networks, whereas the majority have few. Next, a Topological Overlap Matrix (TOM) is created using the adjacency matrix (see Eq. 3). Both direct and indirect gene interactions are reflected in TOM. Based on their shared neighbors, it calculates the degree of connection between two genes.

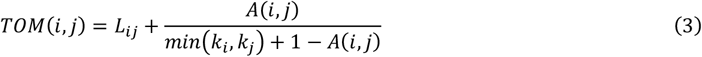

where *L*_*ij*_ is the number of nodes to which both *i* and *j* are connected, and *k*_*i*_ and *k*_*j*_ are the connectivity of node *i* and *j*. Hence, TOM is used to get the measure of dissimilarity between gene pairs based on their network connectivity.

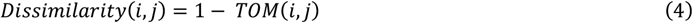

#### 4.2.2. Detection of Gene Modules

Genes with comparable expression characteristics are grouped using hierarchical clustering. A dissimilarity measure calculated from TOM serves as the basis for grouping (see Eq. 4). A hierarchical clustering tree is displayed for each module, which is a collection of highly connected genes. To define the gene modules, a technique known as Dynamic Tree Cut is employed. It facilitates the automatic identification of modules with varying sizes. Compared to traditional methods, this approach is more adaptable. A module eigengene is calculated for every module. The first major element of the gene expression profiles in the module is the eigengene. It symbolizes the module’s general pattern of expression.

#### 4.2.3. Linking modules to external traits

Clustering-derived modules can be associated with external clinical characteristics like phenotype or illness state. This makes it easier to determine whether a module has a strong correlation with any of these characteristics. The module eigengenes (ME) and the clinical features are associated in the module-trait correlation stage. The modules that may be biologically relevant to the trait of interest are revealed by this study. It is beneficial to concentrate on particular gene modules for additional research.

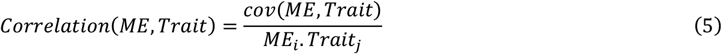

Gene significance, which can be computed for each gene, is a measure of how strongly a particular gene’s expression corresponds with a particular attribute. Furthermore, a gene’s intra-module connectivity indicates how closely it is linked to other genes in the same module. “Hub genes”—genes with significant intra-module connectivity—are thought to be key participants in biological pathways. Hub genes are chosen based on both their gene relevance and intra-module connection. It is believed that these hub genes play a major role in the complicated characteristics or illness being studied.

#### 4.2.4. Hub Gene Identification

Gene significance, which can be computed for each gene, is a measure of how strongly a particular gene’s expression corresponds with a particular attribute. Furthermore, a gene’s intra-module connectivity indicates how closely it is linked to other genes in the same module. “Hub genes”—genes with significant intra-module connectivity—are thought to be key participants in biological pathways. Hub genes are chosen based on both their gene relevance and intra-module connection. It is believed that these hub genes play a major role in the complicated characteristics or illnesses being studied.

#### 4.2.5. Visualization and Validation

Cytoscape is one tool that may be used to visualize the modules and hub genes in the generated networks. Investigating the relationships and organization within the gene network is aided by this. Functional enrichment analysis is used to comprehend the hub genes’ and modules’ biological importance. Common techniques include KEGG pathway analysis and Gene Ontology (GO). The pathways and functions linked to the genes are highlighted in these studies. The WGCNA offers a robust framework for locating gene modules and connecting them to clinical characteristics. Finding promising biomarkers or therapeutic targets of interest is aided by this method.

### 4.3. Analysis of Gene Expression Patterns in HCM through Statistical / AI / ML Approaches

For gene selection in HCM, this part of the study combines statistical and AI/ML techniques. Variance-based screening, volcano plot analysis, and GANGA, a proposed genetic algorithm are used to select genes. To find overlapping genes—possibly in addition to other identified gene sets—the chosen genes from each technique are compared. We build distinct gene regulatory networks (GRNs) for healthy and sick situations using these common genes. By examining the variations in regulatory dynamics between the two states, this method enables us to gain an understanding of the molecular processes that underlie HCM.

#### 4.3.1. Gene Selection

Understanding genetics requires the ability to identify disease-causing genes from gene expression data. Finding a subset of genes that show notable variations in expression between healthy and HCM-affected samples is the goal of gene selection techniques here. Statistical and machine learning approaches that rank genes according to factors including variance, fold-change, and statistical significance (e.g., utilizing volcano plots) are frequently used in gene selection. By maximizing the variations between healthy and diseased profiles, sophisticated techniques such as genetic algorithms further optimize this selection. The chosen genes offer useful targets for comprehending the underlying causes of the disease and possible treatments for HCM. Below is a summary of the gene selection techniques used in our investigation. Below, we outline several gene selection techniques that were used in our investigation.

A. <u>Variance-based Gene Selection</u>: Variance-based gene selection is a method for identifying which genes exhibit the greatest variation in expression levels across different samples. This method calculates the variance of each gene’s expression levels, which shows how much each gene’s expression varies between samples. Since they are more likely to distinguish between different conditions, such as normal and diseased states, genes with higher variance are given priority. Here, Eq. 6 is used to determine variance.

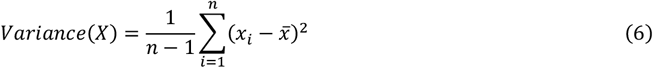
B. <u>Volcano Plot</u>: A volcano plot visually represents the extent of changes in gene expression to their statistical significance by combining fold-change and p-value data. Because of their large fold-change and low p-values, the genes in the plot’s outer regions are prime candidates for further study. Through this method, a specific set of statistically significant and significantly differentially expressed genes can be quickly identified for additional study. Fold change (FC) and P-value, which are related to the plot’s x and y axes, are the two key terms here. FC, which is frequently expressed on a log2 scale (determined by Eq. 7), is the ratio of the mean expression levels of a gene between two states (e.g., diseased vs. normal).

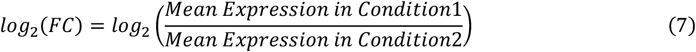
C. <u>Genetic Algorithm for Novel Gene Acquisition (GANGA)</u>: A genetic algorithm (GA) is an optimization method that solves complicated problems by simulating the course of natural development (Holland, 1975). It starts by initializing a population of potential solutions, with a distinct set of values (chromosomes) representing each individual. The algorithm assesses each person’s fitness according to a predetermined fitness function that gauges how successfully they complete the task. Individuals with greater fitness for reproduction are probabilistically selected through the selection process, which frequently employs techniques like roulette wheel selection. Offspring are then produced by applying genetic procedures like crossover and mutation. While mutation introduces random changes to preserve population variety, crossover combines elements of two parent solutions to generate new individuals. Elitism is frequently employed to ensure that the best people’s positive attributes are passed down through the generations. In order to maximize the fitness score, the algorithm constantly evolves the population by iterating over multiple generations. When a stopping criterion—like a predetermined number of generations or fitness convergence—is satisfied, the process comes to an end and the best solution is given back as the optimal or nearly optimal outcome.

In order to find a subset of genes that best distinguish between healthy and pathological states, a customized genetic algorithm, GANGA is developed in this work. A possible gene subset is shown by each chromosome. GANGA optimizes the absolute mean difference in the expression levels of certain genes between the two conditions using a physiologically driven objective function (described in Eq. 8). GANGA uses specific techniques in selection, two-point crossover, and mutation to improve performance while maintaining variety and preventing gene repetition within individuals. Gene subsets are refined by GANGA across several generations, resulting in physiologically significant selections that capture molecular signals and disease-specific expression patterns.

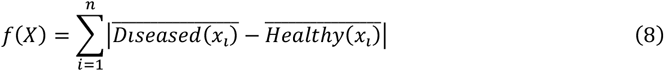

The steps of the algorithm (see Algorithm 1), comprising special features in selection, crossover, and mutation, are as depicted below:

- *<u>Selection</u>*: In GANGA, roulette wheel selection is a probabilistic technique that chooses individuals for reproduction according to their fitness. Similar to how different parts of a roulette wheel vary in size, each person’s likelihood of getting chosen is proportionate to how fit they are with the population as a whole. To retain genetic variety in the population, less fit individuals are still allowed to contribute, while more fit individuals occupy greater segments of the wheel, increasing their possibility of selection. The Roulette Wheel Selection probability for each individual in this GA implementation is determined by comparing its fitness score to the population’s overall fitness. The formula (see Eq. 9) provides the Selection Probability of an Individual (SPI).

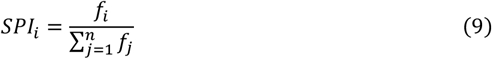
- *<u>Crossover</u>*: A genetic operator called crossover is utilized to create new children by combining the genetic material of two parent solutions. By introducing diversity into the population and simulating biological reproduction, it allows the algorithm to explore new areas of the solution space. This application combines the genetic material of two parents to create new offspring via a two-point crossover approach. The gene symbols in the chromosome have two randomly chosen crossing sites. The crossover scheme is referred to as Two Point Crossover. Parent 1 genes are taken up to the first point for Child 1, then Parent 2 genes in between the points, and finally the remaining Parent 1 genes. Child 2 is similarly developed, with the parents’ responsibilities reversed. This strategy promotes genetic variety in the population by guaranteeing that both offspring inherit portions from each parent.
- *<u>Mutation</u>*: A genetic operator known as mutation causes arbitrary alterations to certain genes within a solution, which effectively alters a gene symbol in our designated candidate solution, titled chromosome. It allows the algorithm to investigate new and maybe better solutions, preventing premature convergence and preserving genetic variety within the population. Here, a parameter called *mutation*_*rate*, determines the probability that the mutation process will occur. Customizing *mutation*_*rate* and few considerations that have been made regarding mutation are as follows:
  – Customizing *mutation*_*rate*: The *mutation*_*rate* is set to 0.1 (10%), which indicates that there is a 10% probability of mutation for every individual.
  – Gene Replacement Strategy: A gene pool is first created from all potential genes in the dataset if a chromosome is chosen for mutation. In order to identify genes that are available for mutation, the genes that are already present in the chromosome are then identified and removed from the pool. Out of this accessible set, a new gene is chosen at random. In order to prevent duplication and preserve individual variability, the new gene is then inserted into the chromosome in place of a randomly selected gene.
  – Ensuring No Repetition: By substituting a gene that is not previously present in the chromosome, the mutation stage guarantees that every gene within a person stays distinct. By doing this, duplication is avoided and each gene subset’s uniqueness is maintained. The algorithm preserves genetic diversity across the population and lowers the possibility of early convergence to less-than-ideal solutions by introducing new genes during mutation, encouraging a more thorough investigation of the solution space.

##### Algorithm 1

Workflow of the GANGA algorithm. Population initialization, fitness assessment using a disease-specific goal, elitism-based selection, two-point crossover, and non-repetitive mutation are all steps in the process. The best gene subsets that differentiate between healthy and sick states are produced through iterative refinement.

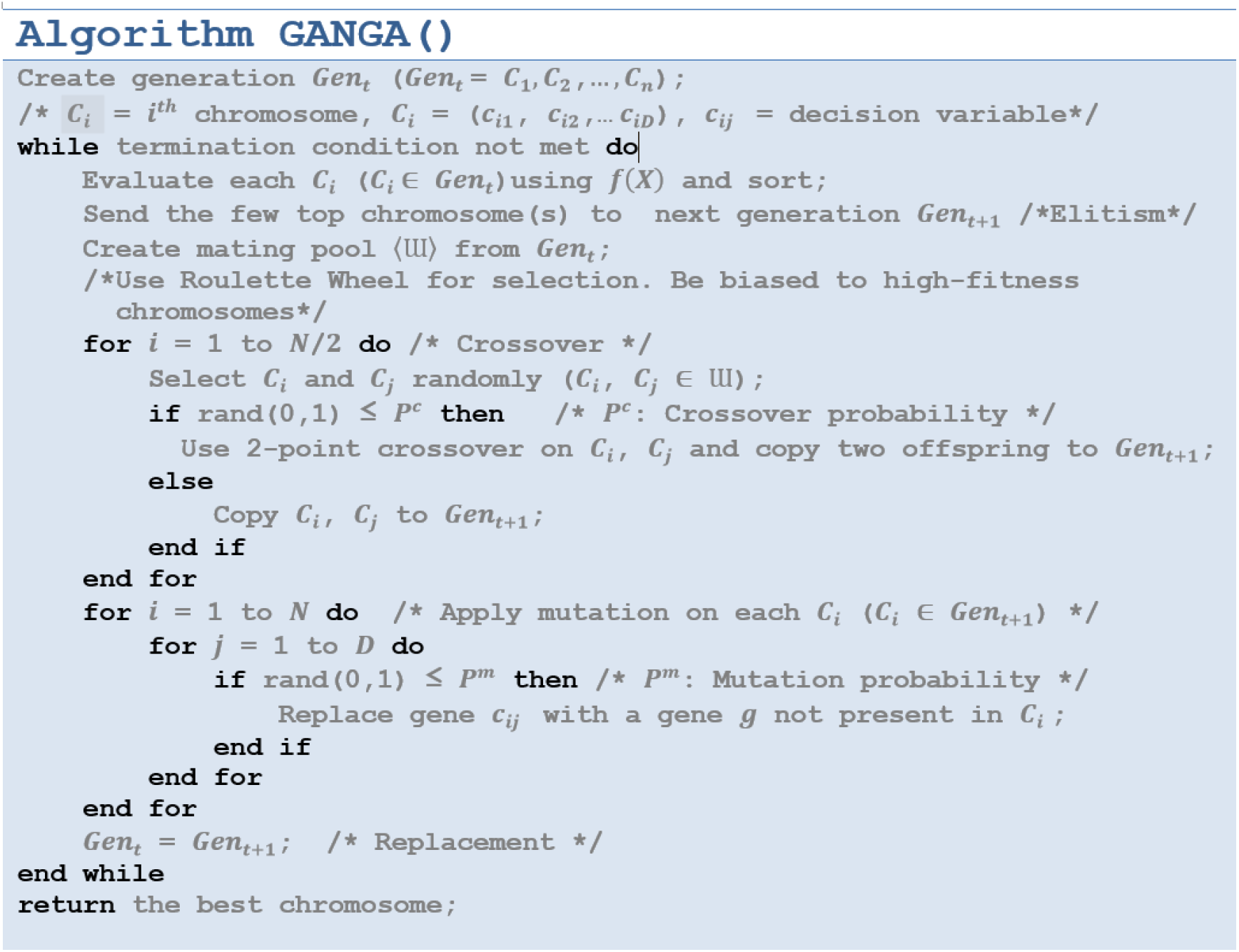

<u>D. Finding common genes across different methods</u>: To identify the most relevant genes, an overlap analysis was performed, where the common genes obtained from these different methods were identified. This approach helps to ensure that the selected genes are consistently significant across various statistical and machine learning techniques, increasing their reliability as potential biomarkers.

#### 4.3.2. Inference of Differential Networks of HCM

Correlation analysis and thresholding approaches are used in this section to infer Differential Networks in healthy and diseased states. Correlation coefficients between the selected genes were first determined by processing the gene expression data from both healthy and sick samples. The strength of the relationships between the genes’ coexpression linkages is shown by these coefficients. To ensure that only the strongest connections were retained for further analysis, a thresholding step was employed to exclude weak correlations from the correlation matrix.

### 4.4. Network Analysis

In order to find notable variations in gene relationships, differential network analysis compares gene co-expression networks from various circumstances, such as diseased and normal states. The significance of each gene inside these networks is evaluated in this study using centrality measures. Key regulatory genes that are essential to the structure and operation of the network can be found using centrality metrics including degree centrality, betweenness centrality, and proximity centrality. By looking at these centrality metrics, this study uses centrality analysis on the differential (healthy and diseased) correlation networks of HCM to find hub genes that significantly influence the molecular mechanisms of HCM. This technique aids in locating potential biomarkers and treatment targets for further investigation.

Metascape, an integrated platform for biological interpretation and functional annotation, was used to validate PPI (Protein–Protein Interaction) and perform enrichment analysis. The study’s gene sets were uploaded to Metascape in order to find biological processes, pathways (such KEGG and GO), and illness connections that were significantly enriched. Metascape’s PPI analysis module, which incorporates information from databases such as BioGrid, InWeb_IM, and OmniPath, was utilized for network-level validation. Confirming the biological significance of the chosen genes and their roles in disease-related pathways, the generated PPI networks showed densely coupled modules that represented functionally coherent gene clusters.

## 5. RESULTS

The outcomes of applying the suggested analytical approach to HCM gene expression data are described in detail in this section. WGCNA, variance, volcano plot, and the GANGA algorithm were used to identify significant gene subsets, and their overlaps were then examined. For both healthy and ill samples, co-expression networks were built independently, and topological alterations were investigated. To find important regulatory genes that might be responsible for the change from healthy to pathological states, centrality analysis on the differential co-expression networks were also carried out. Metascape was used to perform enrichment and protein–protein interaction (PPI) studies in order to comprehend the functional significance of the chosen genes.

### 5.1. Data Source and Preprocessing Outcomes

The dataset GSE36961, available through the GEO repository (NCBI Accession ID), consists of gene expression profiles obtained from a case-control study focused on HCM. The dataset includes transcriptome data from cardiac tissues of patients with HCM, compared to control donor cardiac tissues. Using high-throughput gene expression profiling technology, this dataset provides a comprehensive view of the mRNA transcriptome, offering valuable insights into the molecular mechanisms involved in HCM. The data has been generated from human cardiac tissue samples (Homo sapiens) and serves as a critical resource for further research into the genetic and molecular factors contributing to the development and progression of HCM. The data contains 145 patients and 37846 genes obtained from the surgical myectomy tissue from patients with HCM and normal humans. Details of the data are tabulated in Table 1.

**Table 1.** Details of gene expression data (Geo accession number GSE36961) of surgical myectomy tissue from patients with HCM and normal.

| Features | Healthy/ Control | HCM Patients |
| --- | --- | --- |
| Age | Between 6-75 | Between 9-78 |
| Male | 19 | 54 |
| Female | 20 | 52 |
| Total | 39 | 106 |

### 5.2. Results and Insights from WGCNA Analysis

Using HCM gene expression data obtained from NCBI, the WGCNA method was used to examine differentiability in HCM patients and healthy controls. Different programs/utilities under WGCNA were used. The expression matrix was extracted using the *phenoData*() program. The correlations between gene pairs were measured using a similarity matrix. Next, we created a weighted adjacency matrix by applying soft-thresholding using the *pickSoftThreshold*() function. Similarity and dissimilarity matrices were calculated using topological similarity measurements. In order to create a gene dendrogram, hierarchical clustering was used. *CutreeDynamic*() was used to identify modules, and modules that had at least 75% of similarities were combined. Each module was summarized by computing its eigengenes which is the first principal component of a gene module and represents its overall expression pattern.

Finally, we get two outputs fromThe module dendogram is a tree-like visualization used to identify and group genes that exhibit correlated expression patterns A dendrogram based on eigengene dissimilarities was produced, and inter-module linkages were evaluated using Pearson correlation between eigengenes. the above-mentioned WGCNA method as below:

- We can correlate (Pearson correlation) the found modules’ eigengenes with the particular features (in our case, “disease state”) provided by the phenotypic data. This shows how the module or modules and the trait or traits are related.

- By using Pearson correlation between the genes and their module (intra-modular correlation), we can ultimately find driver genes from identified modules. Module memberships are identified in this way. The driving genes are represented by a gene’s higher module memberships.

For building gene co-expression networks in the HCM dataset using WGCNA, a soft-thresholding power of 10 to 20 strikes a decent mix between scale-free topology (fit index = 0.9) and mean connectedness (see Supplement 1). Genes with associated expression patterns are identified and grouped using the module dendogram, is depicted in Figure 2. In order to find modules that may be related to disease mechanisms, the module-trait heatmap is commonly used to illustrate the relationship between module eigengenes and clinical traits. Such heatmaps can help direct the identification of gene modules linked to disease in the context of HCM (see supplement 2). Table 2 lists the top 25 significant genes (Hub genes) with the strongest intramodular connectivity along with the corresponding variance p-values.

**Table 2.**
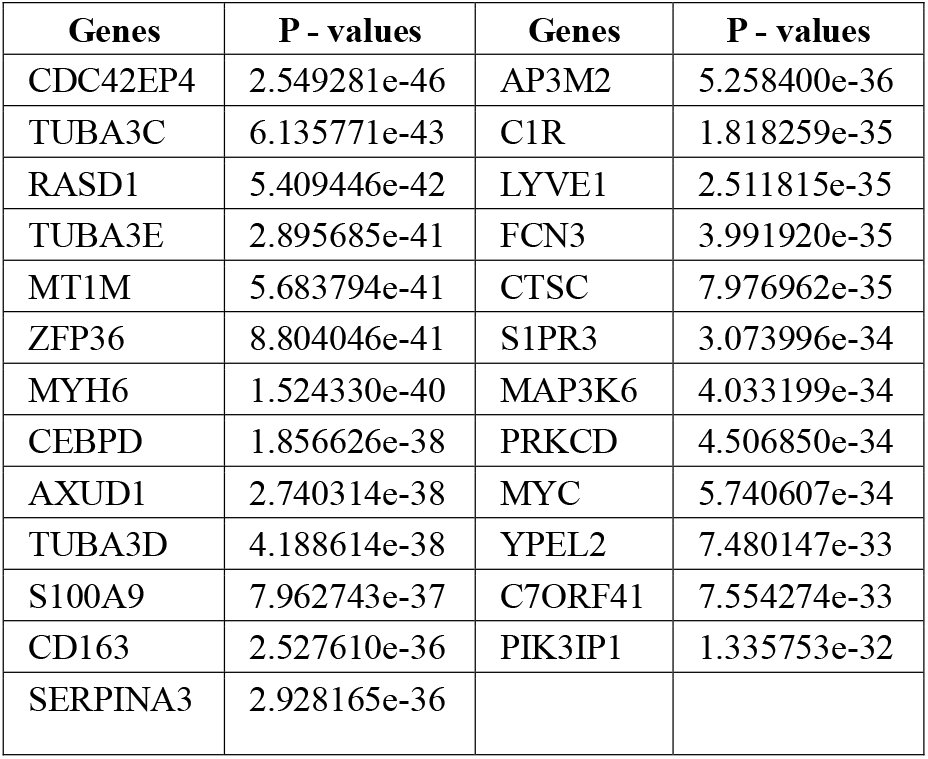
Significant genes identified from WGCNA along with their associated variance p-values.

| Genes | P - values | Genes | P - values |
| --- | --- | --- | --- |
| CDC42EP4 | 2.549281e-46 | AP3M2 | 5.258400e-36 |
| TUBA3C | 6.135771e-43 | C1R | 1.818259e-35 |
| RASD1 | 5.409446e-42 | LYVE1 | 2.511815e-35 |
| TUBA3E | 2.895685e-41 | FCN3 | 3.991920e-35 |
| MT1M | 5.683794e-41 | CTSC | 7.976962e-35 |
| ZFP36 | 8.804046e-41 | S1PR3 | 3.073996e-34 |
| MYH6 | 1.524330e-40 | MAP3K6 | 4.033199e-34 |
| CEBPD | 1.856626e-38 | PRKCD | 4.506850e-34 |
| AXUD1 | 2.740314e-38 | MYC | 5.740607e-34 |
| TUBA3D | 4.188614e-38 | YPEL2 | 7.480147e-33 |
| S100A9 | 7.962743e-37 | C7ORF41 | 7.554274e-33 |
| CD163 | 2.527610e-36 | PIK3IP1 | 1.335753e-32 |
| SERPINA3 | 2.928165e-36 |  |  |

**Figure 2.**
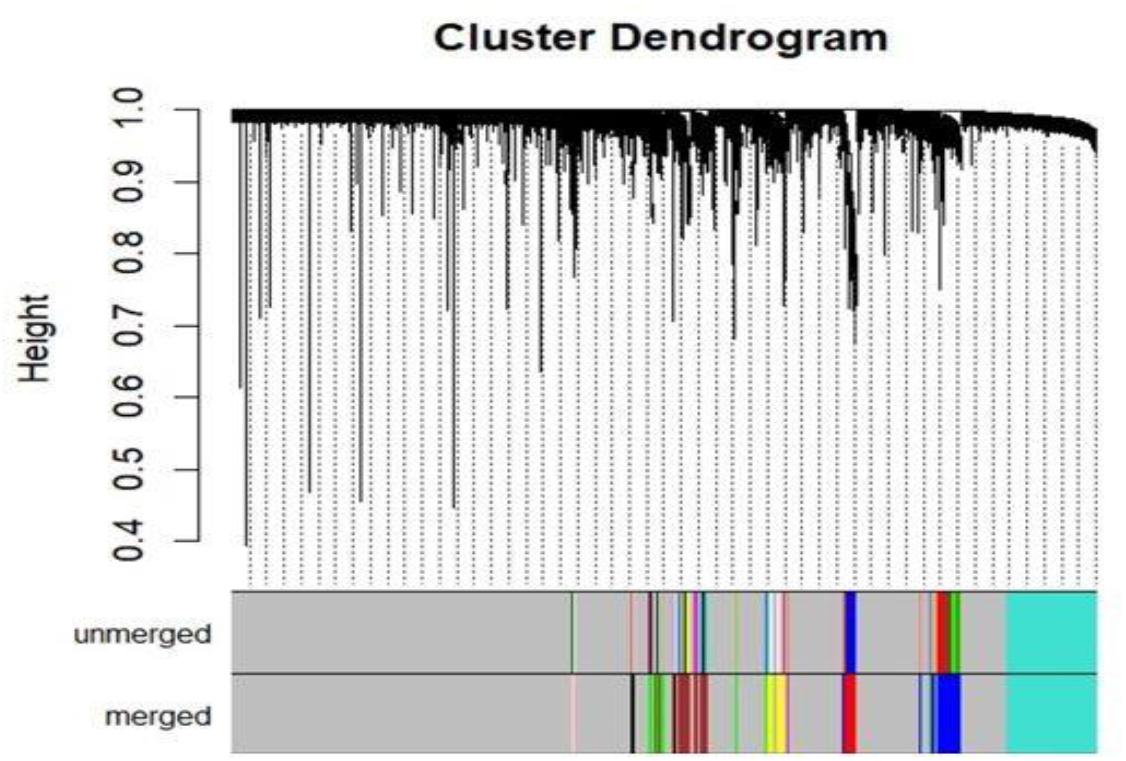
Module dendrogram showing clustering of genes into co-expression modules based on hierarchical clustering using WGCNA.

### 5.3. Results and Insights from STAT/AI/ML Analysis

The AI/ML-based gene selection process also made use of the same dataset, GSE36961, which was previously used for WGCNA analysis. As required by common machine learning methods, the gene expression matrix was reformatted using a transpose operation, which turned genes into columns and samples into rows.

#### 5.3.1. Result and Analysis of Gene Selection

Python 3.0 was used to develop a combination of statistics, artificial intelligence, and machine learning techniques for the analysis of gene expression data. For data preprocessing, statistical calculations, and machine learning model creation, important libraries such as NumPy, pandas, SciPy, and scikit-learn were utilized. These findings provided important insights into the molecular mechanisms behind HCM and facilitated efficient gene selection and co-expression pattern research.

A. <u>Results of Variance Analysis</u>: Critical information about the most variable genes is revealed by the variance analysis of HCM gene expression data, underscoring their possible function in distinguishing HCM from normal states. The top 25 genes (see Table 3) were found by computing the variance of each gene in the differential expression data and then arranging them in descending order. According to the research, genes such as EIF1AY, RPS4Y1, NPPB, SERPINA3, NPPA, RSDT1, and GSTT1 have a significantly high variance, indicating that their expression varies significantly between normal and HCM circumstances (see Figure 3 A). Comprehending the variation in gene expression aids in identifying the molecular processes that underlie HCM.
B. <u>Results of Volcano Plot Analysis</u>: The differential gene expression analysis for HCM is comprehensively visualized by the volcano plot (see Figure 3 B). With the x-axis showing the log2 fold change and the y-axis showing the −log10 of the adjusted p-value, which indicates the statistical significance of changes in gene expression, each point on the figure represents a gene. Significantly elevated genes are plotted in red, and significantly downregulated genes are plotted in blue. 97 genes were found to be elevated, with ACE2, AMHR2, APCDD1L, APOA1, and ATP1B4 being prominent examples. These genes are crucial for processes linked to the development of illness and heart function. On the other hand, 216 downregulated genes were found, such as ADAM11, ADAMTS9, AGXT2L1, and ALDH1A3, which are implicated in a variety of biological activities linked to metabolic pathways and extracellular matrix remodeling. Genes with no discernible changes in expression that did not meet the significance threshold are displayed in gray. An absolute log2 fold change of more than 0.5 and an adjusted p-value of less than 0.05 were the thresholds used to assess significance. Genes that exhibit differential expression in HCM can be easily distinguished thanks to this graphic.
C. <u>Results of GANGA</u>: The customized Genetic Algorithm, GANGA (see Algorithm 1), was applied for gene selection using a population of 50 chromosomes. Each person encoded 40 genes in total. The algorithm had a mutation rate of 0.1 and evolved over 50,000 generations. Each generation’s top performer was retained using an elitism count of 1. These settings were adjusted to guarantee efficient investigation and steady improvement of gene subsets. In the direction of an ideal solution, GANGA stabilized, according to the convergence curve (see Figure 3 C). Between the HCM and control samples, the chosen genes showed a significant expression-level difference. This demonstrates how effective GANGA is in locating genes linked to disease. The final list of important genes derived from GANGA is shown in Table 4, and the next sections provide additional confirmation.
D. <u>Common Genes across Different Algorithms</u>: To examine several gene selection techniques and find important genes linked to HCM, a Venn diagram was employed. Variance-based selection, volcano plot analysis, WGCNA, and a customized genetic algorithm, GANGA were the four techniques used. Ten genes were first found to be shared by the statistical and AI/ML-based approaches (variance, volcano, and GA). These included PDK4, CEBPD, APOA1, MT1X, CENPA, RASD1, S100A8, CORIN, S100A8, and SERPINE1. All four approaches consistently chose three genes (RASD1, CEBPD, and S100A9) when WGCNA was included. These genes may function as biomarkers or therapeutic targets and are probably important in the development of HCM.

**Table 3.** Significant genes from variance analysis.

|  |  |  |  |  |
| --- | --- | --- | --- | --- |
| EIF1AY | RPS4Y1 | NPPB | SERPINA3 | NPPA |
| RASD1 | GSTT1 | HLA-A29.1 | HLA-DRB5 | S100A9 |
| S100A8 | HLA-DRB1 | PDK4 | CORIN | SERPINE1 |
| HMGCS2 | HS.390250 | C6 | MT1X | APOA1 |
| XIST | MYL4 | LOC100008589 | CENPA | CEBPD |

**Figure 3.**
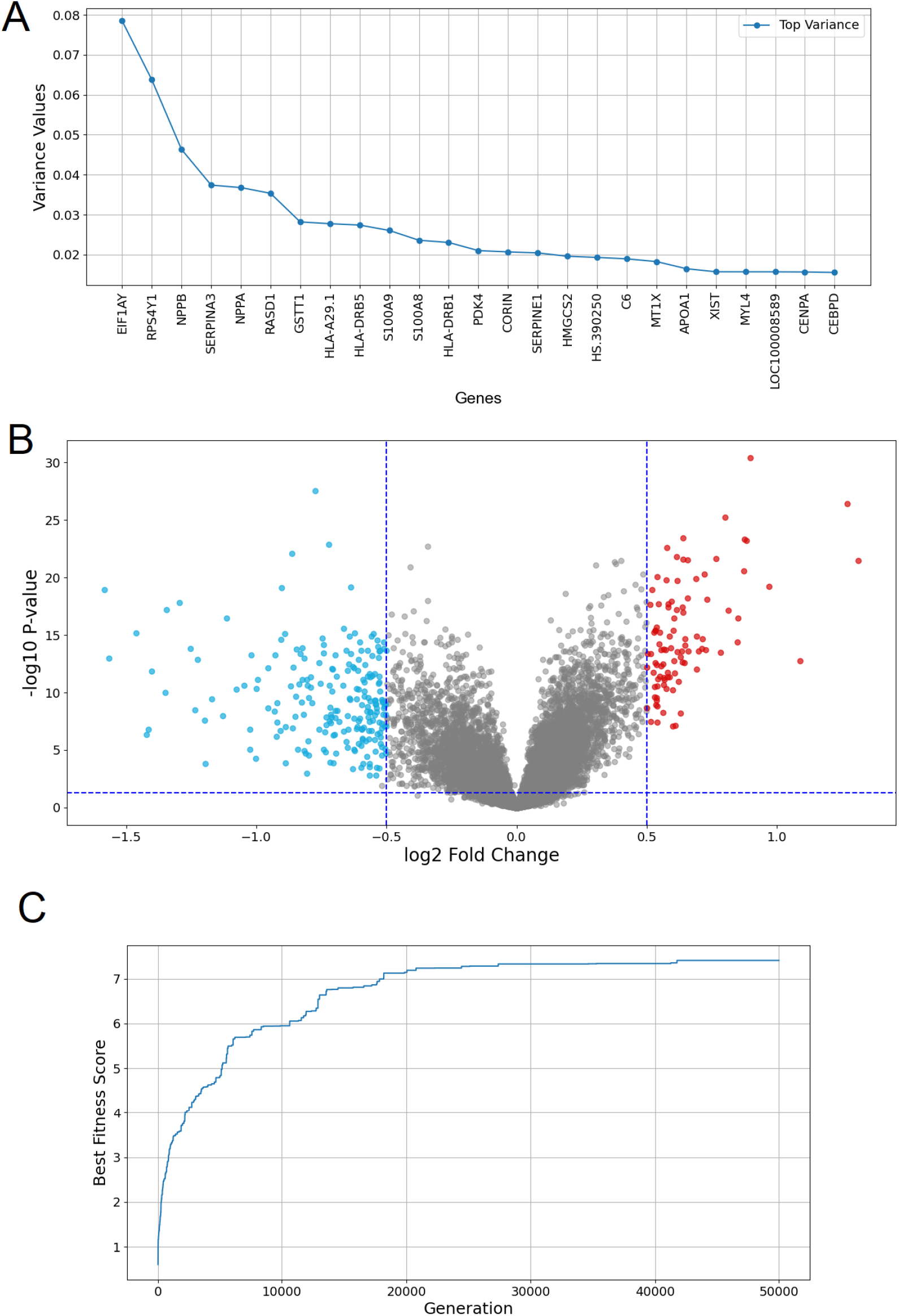
Different statistical/AI/ML methods of gene selection for HCM. (A) Variance analysis of the top 25 genes (B) Volcano plot of differential gene expression analysis (C) Optimization convergence curve of GANGA

**Table 4.** Significant genes obtained from GANGA.

|  |  |  |  |  |
| --- | --- | --- | --- | --- |
| ITGB2 | CD163 | MT1A | C7ORF41 | MYC |
| TUBA3E | FCN3 | TUBA3D | ZFP36 | CORIN |
| HSPA2 | S100A9 | HS.576694 | FKBP5 | RASD1 |
| FCER1G | SERPINE1 | CEBPD | TUBA3C | MT1M |
| RASL11B | SERPINH1 | ELL2 | PIK3IP1 | PDK4 |
| ITGB2 | CD163 | MT1A | C7ORF41 | MYC |
| TUBA3E | FCN3 | TUBA3D | ZFP36 | CORIN |
| HSPA2 | S100A9 | HS.576694 | FKBP5 | RASD1 |

The combined analysis of important metrics from the HCM differential gene expression investigation is displayed in Figure 3. (A) Variance analysis of the top 25 genes in the differential HCM dataset, where variance values are shown on the y-axis and gene names are shown on the x-axis. Genes with high variance, such as EIF1AY, RPS4Y1, NPPB, SERPINA3, NPPA, RSDT1, and GSTT1, were found using techniques like variance-based analysis. B) A volcano plot of HCM differential gene expression analysis utilizing statistical tests like t-tests and adjusted p-values, where the y-axis displays the −log10 adjusted p-value and the x-axis represents the log2 fold change. Notable downregulated genes (blue), such as ADAM11, ADAMTS9, AGXT2L1, and ALDH1A3 (total 216), and upregulated genes (red), such as ACE2, AMHR2, APCDD1L, APOA1, and ATP1B4 (total 97), are indicated. Genes that are not important are displayed in gray. The following thresholds were applied: |log2 fold change| larger than 0.5 and adjusted p-value less than 0.05. (C) The genetic algorithm’s (GA) optimization convergence curve over 50,000 generations shows how the population’s fitness score increases over time. The efficiency of GANGA in fine-tuning gene selection through parameter tuning was demonstrated here.

Panel A of Figure 4 displays the common genes that were obtained using various approaches. Three distinct statistics and AI/ML techniques—variance-based selection, volcano plot analysis, and GANGA— are used in this Venn diagram to depict the gene selection procedure for HCM. SERPINE1, S100A9, CORIN, S100A8, RASD1, PDK4, CEBPD, APOA1, MT1X, and CENPA were found to be ten frequent genes. Three critical genes—RASD1, CEBPD, and S100A9—were identified by an overlap between the three AI/ML approaches when WGCNA was included in the analysis (see Panel B). This suggests that these genes may play a substantial role in the pathophysiology of HCM.

**Figure 4.**
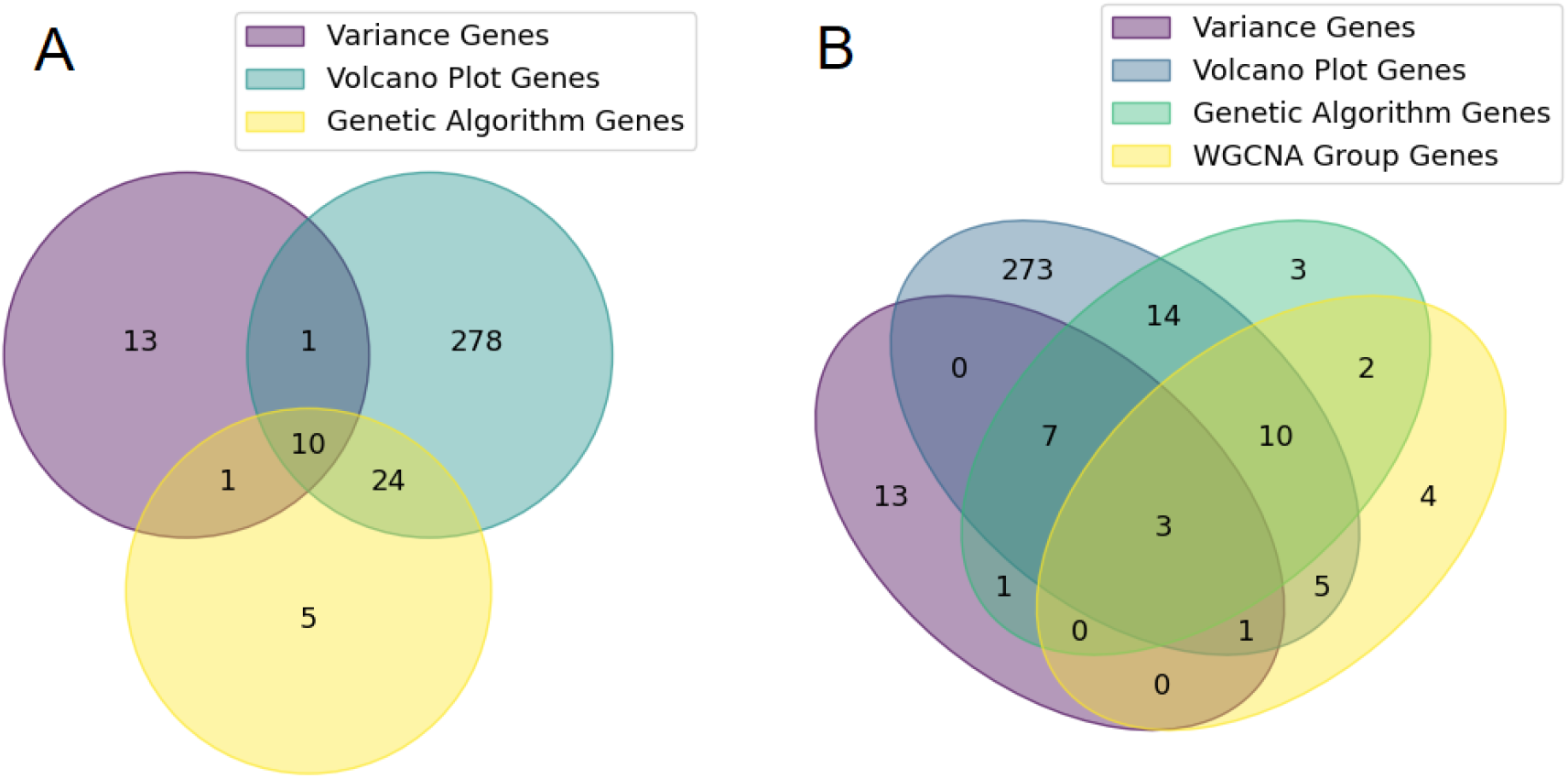
Common HCM disease-critical genes discovered using various techniques. A. Genes that are common to all three statistical, AI, and machine learning techniques. B. Genes that are shared by WGCNA and all statistical, AI, and ML techniques. regulatory network for both health and disease.

#### 5.3.2. Differential Networks of HCM and Centrality Analysis

From the gene expression data of 11 chosen genes in both healthy and HCM states, co-expression networks were deduced. Clear changes in correlation structure and underlying regulatory dynamics were revealed by the considerable changes in gene connectivity between the two circumstances. Figure 5 shows examples of these networks: Panel A displays the baseline regulatory architecture and the co-expression network of healthy individuals. Significant alterations in gene connections were noted in the HCM network, which is displayed in Panel B. A major regulatory gene in the diseased network, S100A9 was crucial in triggering RASD1, which was dormant in the healthy condition. Increased participation of genes like CEBPD and CORIN, which demonstrated high betweenness centrality and were positioned as essential mediators of regulatory flux, was also highlighted by the HCM network design. S100A9 and APOA1, on the other hand, displayed lower degree and closeness centrality in the sick network, suggesting a decline in influence and connectedness. These results highlight changed patterns of gene communication and point to possible treatment targets in HCM.

**Figure 5.**
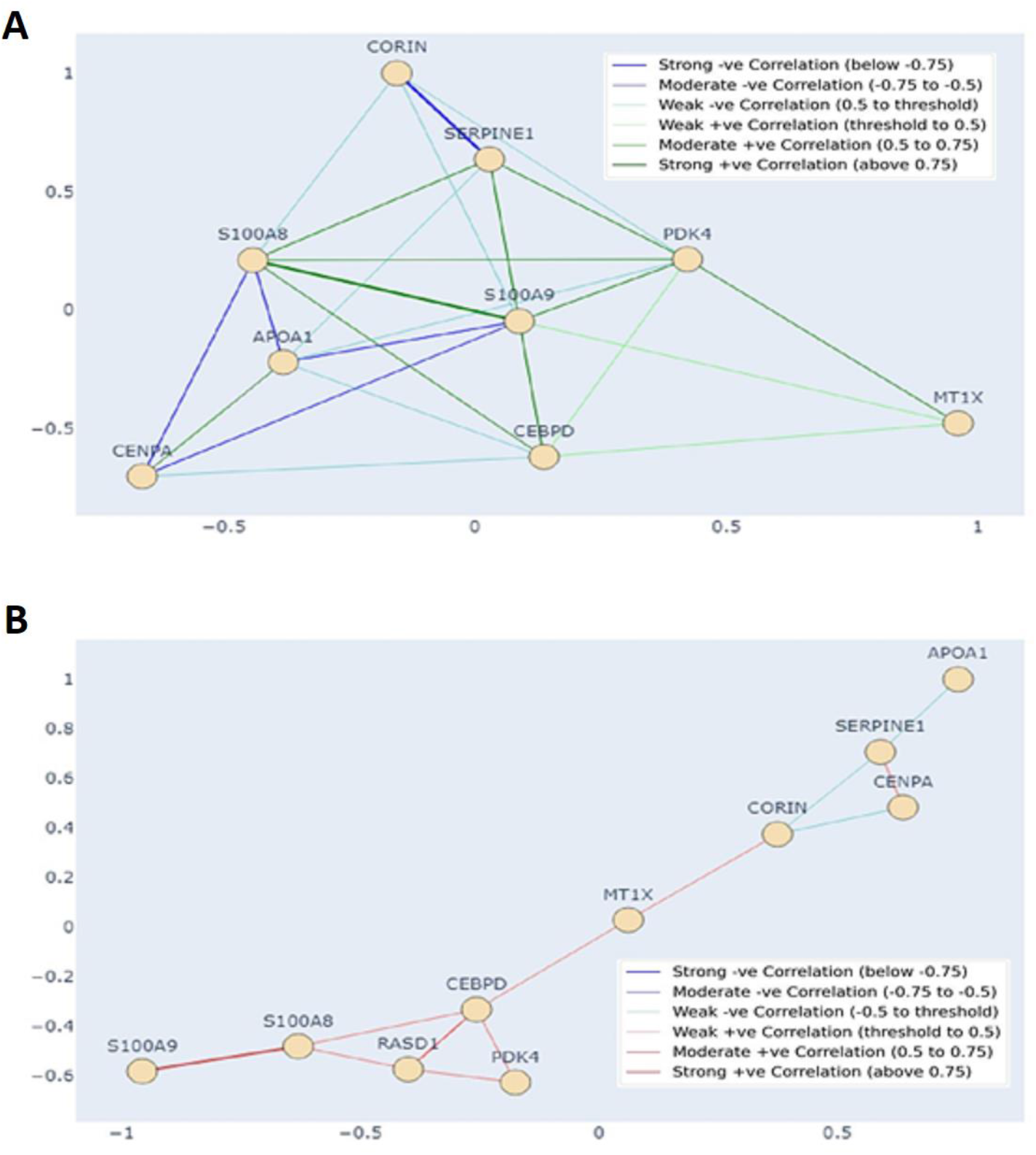
Differential Co-expression Networks of HCM. A. The normal genetic network B. The diseased network

The degree, betweenness, and proximity of centrality measures were calculated to assess each gene’s regulatory impact within the networks. These values are compiled for the 11 chosen genes in Table 5. S100A9 displayed the highest degree and proximity in the healthy network, suggesting robust direct linkage and communication. The network’s structural integrity was supported by the high centrality values of S100A8 and SERPINE1. In the healthy network, RASD1 displayed 0 centrality values, indicating that it played an isolated or dormant role. S100A9, on the other hand, demonstrated more betweenness centrality in the HCM network and served as a crucial link between other genes. Additionally, RASD1, S100A8, and SERPINE1 showed stronger betweenness, indicating that they are more involved in controlling gene interactions in disease. In contrast to the healthy condition, these alterations show how gene impact and communication pathways change within the HCM network.

**Table 5.** Key Gene Centrality Measures in Healthy vs. Disease GRN. Selected genes’ degree, closeness, and betweenness centrality values are shown in this table, emphasizing their interconnectedness and mediating functions within the gene

| Gene Symbols | Healthy |  |  | HCM |  |  |
| --- | --- | --- | --- | --- | --- | --- |
|  | Degree Centrality | Closeness Centrality | Betweenness Centrality | Degree Centrality | Closeness Centrality | Betweenness Centrality |
| SERPINE1 | 0.625 | 0.727 | 0.009 | 0.333 | 0.36 | 0.222 |
| S100A9 | 1 | 1 | 0.146 | 0.111 | 0.29 | 0 |
| S100A8 | 0.875 | 0.889 | 0.089 | 0.333 | 0.391 | 0.222 |
| CORIN | 0.5 | 0.667 | 0 | 0.333 | 0.4 | 0.1 |
| RASD1 | 0 | 0 | 0 | 0.333 | 0.391 | 0.028 |
| PDK4 | 0.875 | 0.889 | 0.089 | 0.722 | 0.36 | 0 |
| CEBPD | 0.75 | 0.8 | 0.01 | 0.444 | 0.5 | 0.383 |
| APOA1 | 0.75 | 0.8 | 0.03 | 0.111 | 0.727 | 0 |
| MT1X | 0.375 | 0.615 | 0 | 0.222 | 0.5 | 0.556 |
| CENPA | 0.5 | 0.667 | 0 | 0.222 | 0.346 | 0 |

### 5.4. Integrated Pathway Enrichment and Interaction Network of Key Genes

For biological validation, key genes found using WGCNA, statistical, and AI/ML-based methods were put through a thorough bioinformatics pipeline. With the complete genome as backdrop, pathway and process enrichment was carried out utilizing seven ontology databases (KEGG, GO Biological Processes, Reactome, CORUM, WikiPathways, PANTHER, and Canonical Pathways). Kappa similarity scores (> 0.3) were used to cluster significant phrases (p < 0.01, enrichment factor > 1.5, minimum 3 genes), with the most significant term from each cluster serving as its representative. Using hypergeometric testing with Benjamini-Hochberg correction, statistical rigor was guaranteed. The most enriched route was found to be cytokine signaling, with node color signifying cluster identity and p-value significance, when a selection of the top 250 keywords (≤15 per cluster) were shown in Cytoscape.

- Disease Association: As seen in Panel A of Figure 6, DisGeNET confirmed the gene set’s clinical significance in myocardial disease.
- Network Topology: PPI modules highlighted how crucial cytokine signaling is for function (see Panel B of Figure 6).
- Pathway Convergence: Coordinated immune-inflammatory activity was emphasized by enrichment clustering.

**Figure 6.**
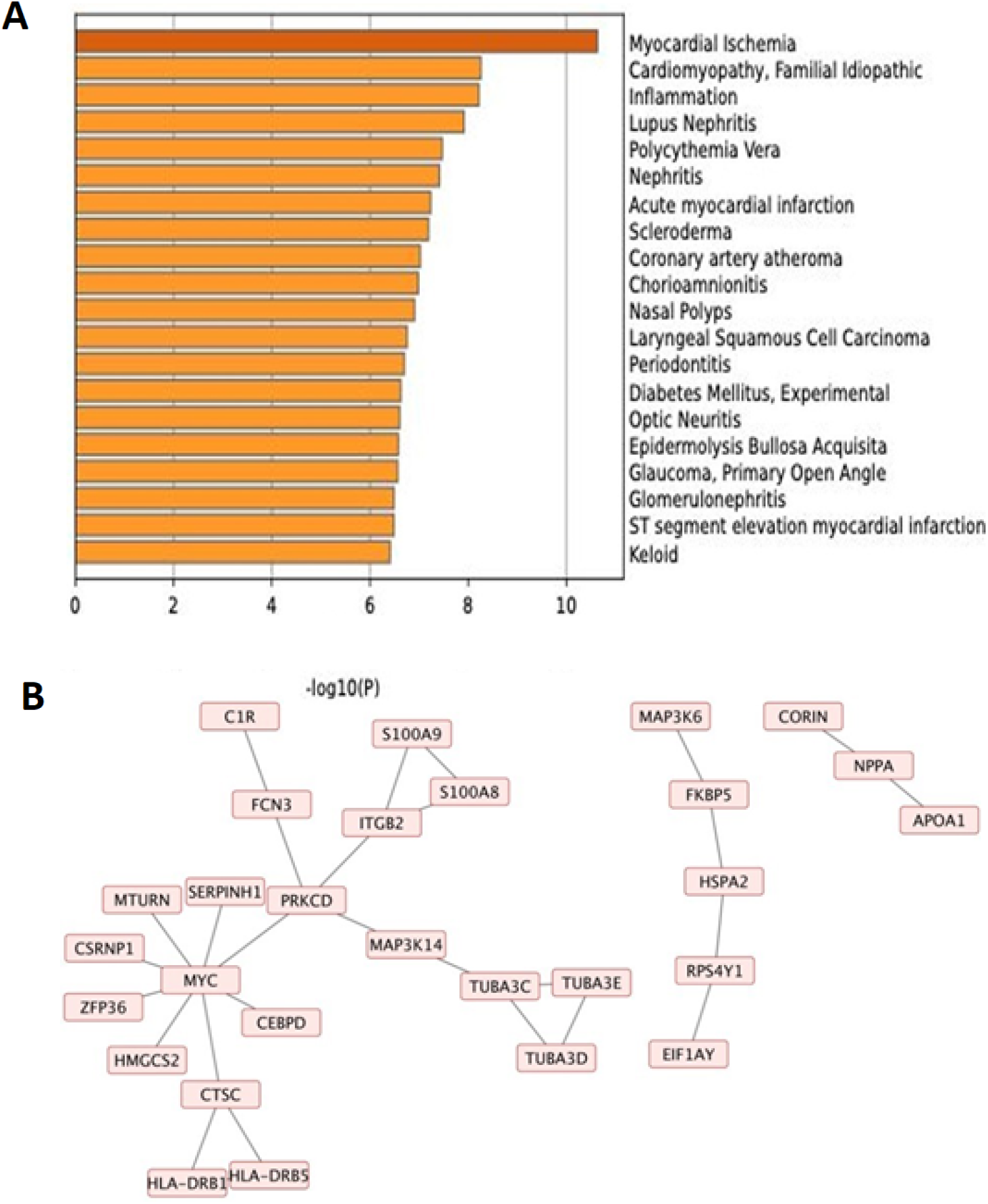
Functional enrichment and PPI network of selected genes. A) Cytokine signaling was identified as the most prominent pathway by enrichment analysis using seven ontology databases (p < 0.01, enrichment factor > 1.5). (B) The PPI network made up of STRING, BioGRID, OmniPath, and InWeb_IM emphasizes modules linked to phagosome activity, interferon response, and cytokine signaling, highlighting the role of inflammatory and immunological pathways in HCM

S100A8/A9, key members of the S100 calcium-binding protein family secreted by myeloid cells, mediate cardiovascular pathophysiology through TLR4 and RAGE signaling, driving inflammatory responses, fibrosis, and vascular dysfunction. In HCM, the S100A8/A9 heterodimer promotes disease progression by activating TLR4/NF*k* B-mediated inflammation inducing IL-6/TNF-α release and leukocyte infiltration (Marinković et al., 2020), enhancing TGF-β-driven fibrosis (Parichatikanond et al., 2020), and disrupting Calcium handling via RAGEERK/p53 pathways, leading to arrhythmogenicity and cardiomyocyte hypertrophy (Yuan et al., 2024c). Clinically, elevated

S100A8/A9 correlates with LV wall thickening, fibrosis burden, and sudden cardiac death risk (Eggers et al., 2011). Experimental studies demonstrate that pressure overload upregulates S100A8/A9, which exacerbates hypertrophy through FGF23-mediated calcineurin/NFAT activation, while genetic deficiency attenuates this response (Bayes-Genis et al., 2020; Chen et al., 2024), highlighting its potential as a therapeutic target in HCM.

## 6. CONCLUSION

The complex cardiac condition known as HCM has a variety of hereditary causes. This study used a multi-step computational methodology to analyze gene expression data in order to discover important genes and regulatory patterns linked to HCM. Using three gene selection methods—variance-based filtering, volcano plot analysis, and a new GANGA—we started by gathering transcriptome data from public repositories. GANGA was created especially to maximize the expression discrepancies between healthy and sick samples in order to optimize gene selection. In order to verify the overlapping genes from all gene selection techniques, PPI analysis examined significant interactions between their encoded proteins, and enrichment analysis evaluated their participation in established biological pathways. We used the gene expression data of the verified common genes in both healthy and HCM states to build differential co-expression networks in order to investigate regulatory alterations. The main regulators causing network reconfiguration in the illness state were then found using centrality analysis.

For gene selection tasks in disease situations, GANGA presents a tailored genetic algorithm framework with targeted alterations. GANGA guarantees diversity and relevance in chosen gene sets by implementing physiologically based tactics like elitism, non-redundant gene encoding, and customized crossover. Its usefulness in identifying disease-specific molecular signatures is demonstrated by its consistent ability to extract genes with high discriminative power between HCM and control samples.

After removing WGCNA findings, our analysis identified eleven common genes, including CORIN, S100A9, and SERPINE1. Three overlapping genes—RASD1, CEBPD, and S100A9—were found when WGCNA was added. The discovered genes’ functional relevance was validated by enrichment and network-based validation. They formed important modules within cytokine-centered protein-protein interaction networks and had strong correlations with pathways related to inflammation. Additional information about the gene interactions and their changes under HCM circumstances was obtained by building differential co-expression networks for both healthy and sick states. In the sick network, S100A9 was found to be a crucial regulatory role, particularly triggering RASD1 from its dormant state, indicating a possible therapeutic target for additional study.

There are a few drawbacks to this study, though, that should be noted. The dependence on current gene expression data, which might not adequately represent the complexity of the genetic and epigenetic components involved in HCM, is one significant drawback. Furthermore, the generalizability of our findings may be impacted by the very small sample size. Additionally, because the analysis was mainly concerned with gene-level data, it might have missed crucial regulatory components such post-translational modifications and non-coding RNAs.

Future studies should try to confirm these results with bigger and more varied cohorts, combining proteomics and metabolomics data for a more thorough comprehension of HCM. Through experimental research, the identified genes—especially S100A9 and RASD1—should be functionally validated. Furthermore, extending the application of cutting-edge machine learning methods may make it easier to create predictive models for HCM risk evaluation. Overall, this study’s findings offer a strong basis for further research aimed at deciphering the intricate genetic pathways underlying HCM, which will ultimately guide the development of innovative treatment approaches and enhance patient outcomes.

## Data Availability

The study used (or will use) ONLY openly available human data that were originally located at:https://www.ncbi.nlm.nih.gov/geo/query/acc.cgi?acc=GSE36961

## Notes

### Competing Interest Statement

The authors have declared no competing interest.

### Funding Statement

No funding

